# Diffusion Histology Imaging to Improve Lesion Detection and Classification in Multiple Sclerosis

**DOI:** 10.1101/19009126

**Authors:** Zezhong Ye, Ajit George, Anthony T. Wu, Xuan Niu, Joshua Lin, Gautam Adusumilli, Anne H. Cross, Peng Sun, Sheng-Kwei Song

## Abstract

**Background:** Diagnosing MS through magnetic resonance imaging (MRI) requires extensive clinical experience and tedious work. Furthermore, MRI-indicated MS lesion locations rarely align with the patients’ symptoms and often contradict with pathology studies. Our lab has developed and modified a novel diffusion basis spectrum imaging (DBSI) technique to address the shortcomings of MRI-based MS diagnoses. Although primary DBSI metrics have been demonstrated to be associated with axonal injury/loss, demyelination and inflammation, a more detailed analysis using multiple DBSI-structural metrics to improve the accuracy of MS lesion detection and differentiation is still needed. Here we report that Diffusion Histology Imaging (DHI), an improved approach that combines a deep neural network (DNN) algorithm with improved DBSI analyses, accurately detected and classified various MS lesion types.

**Methods:** Thirty-eight multiple sclerosis patients were scanned with T2-weighted imaging (T2WI) using fluid attenuated inversion recovery (FLAIR), T1-weighted imaging (T1WI) using magnetization-prepared rapid acquisition with gradient echo (MPRAGE), magnetization transfer contrast (MTR) imaging and diffusion-weighted imaging. The imaging results identified 43,261 voxels from 91 persistent black hole (PBH) lesions, 89 persistent gray hole (PGH) lesions, 16 acute gray hole (AGH) lesions, 189 non-black hole (NBH) lesions and 113 normal-appearing white matter (NAWM) areas. Data extracted from these lesions were randomly split into training, validation, and testing groups with an 8:1:1 ratio. The DNN was constructed with 10 fully-connected hidden layers using TensorFlow 2.0 in Python. Batch normalization and dropout regularization were used for model optimization.

**Results:** Each MS lesion type had unique DBSI derived diffusion metric profiles. Based on these DBSI diffusion metric profiles, DHI achieved a 93.6% overall concordance with neurologist determinations of all five MS lesions, compared with 74.3% from conventional MRI (cMRI)-DNN model, 78.2% from MTR-DNN model, and 80% from DTI-DNN model. DHI also achieved greater performances on detecting individual MS lesion types compared to other models. Specifically, DHI showed great performances on prediction of PBH (AUC: 0.991; F_1_-score: 0.923), PGH (AUC: 0.977; F_1_-score: 0.823) and AGH (AUC: 0.987; F_1_-score: 0.887), which significantly outperformed other models.

**Conclusions:** DHI significantly improves the detection and classification accuracy for various MS lesion types, which could greatly aid the clinical decisions of neurologists and neuroradiologists. The efficacy and efficiency of this DNN model shows great promise for clinical application.

## Introduction

Multiple sclerosis (MS) is a common inflammatory CNS disorder that affects around 900,000 people in the United States^1^. MS usually begins with patients having intermittent “attacks” (i.e. relapsing-remitting course) characterized by transient symptoms of focal central nervous system (CNS) dysfunction, such as numbness or weakness of a limb, incoordination, vertigo or visual dysfunction^2^. These clinical attacks, or relapses, are caused by focal inflammation in the CNS directed against the myelin insulation around the axons^3^. The inflammation results in focal “MS lesions” characterized by demyelination, axonal injury and axonal conduction blockage - all hallmarks of MS^3^.

Identifying and segmenting the focal white matter (WM) lesions is the first essential step in characterizing the disease burden of an MS patient, especially in calculating and interpreting more specific measures of damage. Conventional magnetic resonance imaging (cMRI) is often used to characterize and quantify MS lesions in the brain^4^, where the lesions are classified based on their intensity under different MR sequences^5^. The classical appearance of WM lesions in cMRI manifests as hyperintense regions in T2-weighted images (T2WI) and hypointense region in T1-weighted images (T1WI)^5^.

While standard cMRI is sensitive in detecting various MS lesions, it lacks pathological specificity and requires extensive clinical experience and large workload^6^. Accurate identification of MS lesions using cMRI becomes extremely difficult due to lesion variability in location, size and shape; the anatomical variability between subject further complicates diagnoses^7^. In addition, the association of MS lesions with clinical presentation and pathologies may come in discordance.

Our lab has developed a novel diffusion basis spectrum imaging (DBSI)^8,9^ and demonstrated its capability in quantitatively characterizing the pathologies that underlay MRI lesions in MS patients and post-mortem specimens^10,11^. Despite DBSI metrics have been validated to be specifically associated with axonal injury/loss, demyelination and inflammation, the validity and accuracy of applying DBSI in detecting and differentiating MS lesion based on the multiple DBSI-structural metrics have yet to be established. Therefore, we introduced a novel Diffusion Histology Imaging (DHI) approach that combines a deep neural network (DNN) algorithm with improved DBSI analysis and reported its accuracy in detecting and classifying the various MS lesion types.

## Materials and Methods

### Subject

A total of thirty-eight patients with MS were enrolled in this study. The study was approved by the Institutional Review Board of Washington University School of Medicine. Each participant had provided written informed consent.

### Image Acquisition

Patients were imaged on a 3.0T Siemens Trio scanner (Siemens, Erlangen, Germany). T1WI using magnetization-prepared rapid gradient-echo (MPRAGE) scan with an isotropic 1 mm^3^ resolution was used for identification of structural landmarks and as a registration target (TR=2400 ms, TE=3.16 ms, TI=1000 ms, Matrix=256×224, FOV=256×224 mm^2^ Resolution=1×1×1 mm^3^). T2WI using fluid-attenuated inversion recovery (FLAIR) scan was acquired to quantify visible WM lesion volumes (TR=7500 ms, TI=2500 ms, TE=210 ms, TI=2500 ms, Matrix=256×256, FOV= 256×256 mm^2^, Resolution=1×1×1 mm^3^). Magnetization transfer (MT) images were acquired with the following parameters: Repetition Time (TR)=43ms; Echo Time (TE)=11 ms; Flip Angle=30 degrees; Field of View (FOV)=192×256 mm^2^; slice thickness=3 mm; in-plane resolution=1×1 mm^3^; the number of slices=60; total acquisition time=8 min. Magnetization transfer ratio (MTR) maps were calculated pixel-by-pixel using the equation: MTR= (S_off_–S_on_)/S_off_ ×100, where S_on_ and S_off_ were signal intensities with and without saturation pulse. Axial diffusion-weighted images (DWI) covering the whole brain were acquired using a multi-b value diffusion weighting scheme (99 directions, maximum b-value 1500 s/mm^2^) and the following parameters: TR=10,000 ms; TE=120 ms; FOV=256×256 mm^2^; slice thickness=2 mm; in-plane resolution=2×2 mm^3^; number of slices=56; total acquisition time=15 min. Eddy current and motion artifacts of DWI were corrected, then the susceptibility-induced off-resonance field was estimated and corrected.

### MS Lesion Identification

All the lesions were identified by an experienced neurologist with 20 years of clinical experience. Amira 6.0.1 visualization and analysis software (FEI, Hillsboro, OR) was used to quantify intensity values for each hypointense lesion on all scans. Hypointense areas of white matter (WM) on T1WI were defined as “black holes” (BH). Less hypointense BHs were defined as “gray holes” (GH). Note that the lesion intensity assessment requires establishing the baseline intensity of each scan to account for scan-to-scan variations.

The BH and GH that have persisted for at least 12 months are markers of focal tissue injury in MS and are known as “persistent black holes” (PBH) and “persistent gray holes” (PGH), respectively^12^. BHs and GHs that have persisted for less than 12 months are markers for inflammation and are known as “acute black holes” (ABH) and “acute gray holes” (AGH), respectively. Other T2WI hyperintense non-black/gray hole lesions were defined as non-black hole (NBH) T2W lesions. ABH were excluded in this study because the number of identified ABHs were insufficient for model training.

Persistent black holes (PBH, Fig. 1A), persistent gray hole (PGH, Fig. 1B), acute gray hole (AGH, Fig. 1C), T2-weighted hyperintense non-black hole lesion (NBH, Fig 1D) and normal appearing white matter regions (NAWM) were identified with corresponding lesion masks drawn using Amira.

**Figure 1.**
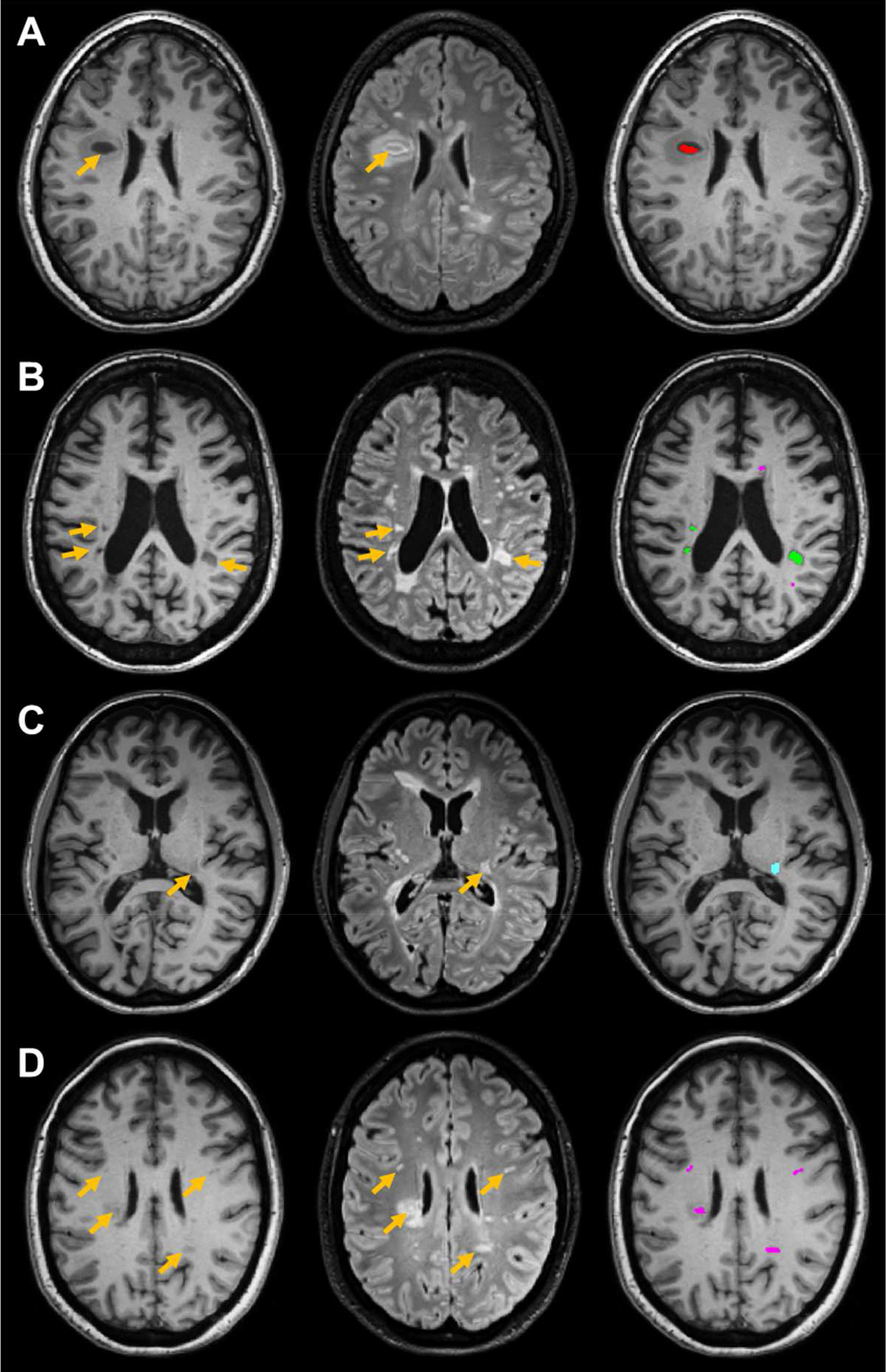
MS lesions as identified by experienced neurologists. Yellow arrows indicate the location of neurologist-identified lesions. Columns contain data from MPRAGE, FLAIR, and MPRAGE with colored markers respectively. **A**. persistent black holes (red). **B**. persistent grey holes (green) with acute grey holes (cyan). **C**. Acute grey holes (cyan). **D**. Non-black hole lesions (purple).

### Diffusion Basis Spectrum Imaging

DBSI metric maps were estimated on pre-processed DW images using an in-house software developed using MATLAB^®^ (Mathwork, MA). DBSI models the diffusion-weighted MRI signals as a linear combination of multiple tensors describing both the discrete anisotropic axonal fibers and an isotropic diffusion spectrum encompassing the full range of diffusivities^9^, as seen in Eq. [1].

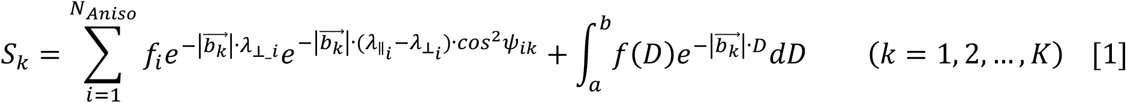

In [1], S_*k*_ and 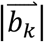 are the normalized signal and b-value of the k^th^ diffusion gradient, *N*_*Aniso*_ is the number of anisotropic tensors, *Ψ*_*ik*_ measures the angle between the kth diffusion gradient and the principal direction of the *i*^*th*^ anisotropic tensor, 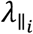 and 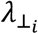 are the axial diffusivity (AD) and radial diffusivity (RD) of the *i*^*th*^ anisotropic tensor, *f*_*i*_ is the signal intensity fraction for the *i*^*th*^ anisotropic tensor, and *a* and *b* are the low and high diffusivity limits for the isotropic diffusion spectrum *f(D)*. The anisotropic diffusion component describes water molecules inside and outside myelinated or non-myelinated axons. DBSI-derived anisotropic signal intensity fractions (*f*_*i*_, i.e., fiber fraction) can also be defined as apparent axonal density in WM. DBSI-derived AD and RD retain the pathological specificity for axonal injury and demyelination as previously proposed models but contain fewer confounds from coexisting MS pathologies. The DBSI-derived “restricted” isotropic diffusion fraction (ADC ≤ 0.3 µm^2^/ms) has been shown to reflect cellularity^9^. Cellular and axonal packing plays a crucial role in extracellular and extra-axonal diffusion characteristics. Less restricted or non-restricted isotropic diffusion components represent water molecules in less densely packed environments, such as areas of tissue disintegration or edema, or non-restricted water within cerebrospinal fluid (CSF) ^9,13,14^.

### Image Processing

Whole-brain voxel-wise DTI and DBSI analyses were performed by an in-house software developed using MATLAB^®^. To control for scan-to-scan variation within individual scans, cerebral spinal fluid (CSF), which is unaffected by MS pathologies, was used as the baseline for individual scans and to asses signal intensities of MS lesions under T1WI and T2WI modalities. Around one hundred representative CSF voxels in the center of the ventricle were selected to ensure the exclusion of any voxels containing choroid plexus or partial volume effect from the ventricular edges. The median intensity was defined as “CSF Intensity” for that scan. For each voxel in MS lesions, the voxel intensity was divided by the CSF intensity to normalize b0, T1WI, and T2WI intensities.

### DNN Model Development and Optimization

Our complete data sample consisted of 43261 imaging voxels from 499 MS lesions coming from 38 patients. The collected voxels were split into training, validation, and testing sets with a ratio of 8:1:1 respectively. We compared four different models, each containing their own set of features. The first model is the DBSI-DNN model (DHI), which used DBSI metrics, normalized T1WI and T2WI intensities; the second model is the conventional MRI (cMRI) DNN model, which used normalized T1WI and T2WI intensities; the third model is the MTR CNN model, which used MTR metrics and normalized T1WI and T2WI intensities; finally, the fourth model is the DTI DNN model, which used DTI metrics and normalized T1WI and T2WI intensities. A total of 16 diffusion metrics from DBSI and 4 DTI metrics were used.

The DNN model was developed using Tensorflow 2.0 frameworks^15^ in Python. DNN models with varying numbers of hidden layers, nodes and training epochs were tested for model optimization. The grid search method was used for the network architecture optimization. Pruning was also adopted to optimize the number of nodes per layer in DNN models. The neural networks were pruned to having between 0 and 200 nodes in each layer in increments of 10 nodes. Pruned neural networks were tested against validation accuracy over 150 epochs. Ten sets of 10-fold cross-validation were performed at unique random states to ensure the statistical relevance of the results. Batch normalization was performed with a mini-batch size of 200 before feeding data to the next hidden layer to improve model optimization and prevent overfitting. Exponential linear units (ELU) were used to activate specific functions in each hidden layer. The final layer was a fully connected softmax layer that produces a likelihood distribution over the five output classes. The network was trained with random initialization of the weights as described in He et al^16^. The Adam optimizer was used with the default parameters of β_1_=0.9 and β_2_=0.999 and a mini-batch size of 200. The learning rate was manually tuned to achieve the fastest convergence (1×10^−3^). The cross-entropy loss function was chosen, and the model was trained to minimize the error rate on the development dataset. The hyper-parameters of the network architecture and optimization algorithm were chosen through a combination of grid search and manual tuning.

### Statistical Analysis

Confusion matrices were calculated and used to illustrate the specific examples of MS lesion classes where the DNN prediction contradicts the neurologist’s diagnoses. The one-versus-rest strategy was implemented to perform ROC analysis and area under curve (AUC) was calculated to assess model discrimination of each lesion type. Sensitivity and specificity values were calculated at the optimal cut off points, which were computed by maximizing the differences between true positive rates and false positive rates. The precision-recall curve was calculated to show the relationship between precision (positive predictive value) and recall (sensitivity/true positive rate) which provides complementary information to the ROC curve when using data with imbalanced classes. F_1_ scores were computed and used to compare the relative performance of the DNN models to the neurologist-identified MS lesions. The F_1_ score favors models that maximize both precision and recall simultaneously, which is especially helpful in the setting of multi-class prediction where the AUC may be insensitive class imbalance. All the 95% confidence interval values were calculated with bootstrap methods with 1000 iterations^17^. Statistical metrics and curves were calculated by packages from Scikit-learn^18^.

## Results

### MS Patient and Lesion Characteristics

A total of 38 patients were recruited for this study. The male to female ratio of the patients was 12 to 26. The patients averaged 55 years old (±10.6 years). Among these patients, 15 had primary progressive MS, 10 had secondary progressive MS and 13 had relapsing remitting MS (Table 1). A neurologist with 20+ years of experience identified regions of interest (ROI), which included 92 PBH lesions, 89 PGH lesions, 16 AGH lesions, 189 NBH lesions and 113 NAWM regions. The average volumes of the MS lesions were 108.3, 66.5, 141.1, 60.6 and 120.9 mm^3^, respectively. Three of the 38 subjects had no PBH or PGH lesions.

**Table 1.** Patient and Lesion characteristics.

| **Patient/Lesion Characteristics** | **No.** |
| --- | --- |
| **Patient** | **38** |
| **Age (years), median (range)** | **55 (25-72)** |
| **Disease duration (years), median (range)** | **13.7 (1.5-43.3)** |
| **EDSS, median (range)** | **6 (1.5-6.5)** |
| **Gender (male/female)** | **12/26** |
| **MS subtypes** |  |
| RRMS | 13 |
| PPMS | 15 |
| SPMS | 10 |
| **MS lesion types** |  |
| PBH | 92 |
| PGH | 89 |
| AGH | 16 |
| NBH | 189 |
| NAWM | 113 |

### Histogram Analysis of Different MRI Metrics

Conventional MRI, DTI and DBSI metrics showed distinguishing distribution profiles among different MS lesion types (Fig. 2). NAWM regions clearly displayed lower normalized FLAIR signals than other lesion types. PBH, PGH, AGH and NBH lesions showed similar distributions with significant overlap. PBH lesions have lower MPRAGE and MTR signals compared to NAWM regions. PBH lesions and PGH lesions also have broader MTR distributions compared to NAWM regions. PBH lesions and PGH lesions have significantly elevated ADC signals of broader distributions compared to NAWM regions, NBH lesions, and AGH lesions. PBH lesions and PGH lesions have significantly narrower DTI-FA distributions. Both PBH and PGH lesions distributions are also shifted towards smaller values when compared to NAWM regions, NBH lesions and AGH lesions. PBH lesions and PGH lesions have slightly narrower fiber fraction distributions compared to NAWM regions, NBH and AGH lesions. Axial and radial diffusivity signals gradually shift towards higher values from NAWM regions, NBH, AGH, PGH to PBH lesions, respectively. While all lesions fall within similar signal isotropic ADC ranges, distinct and complex distribution patterns specific to each type of the lesion show when the DBSI metrics are considered. PBH and PGH lesions have lower restricted fraction signals and much narrower distributions compared to NAWM regions, NBH and AGH lesions. Hindered fraction signals gradually shift towards higher values from NAWM regions, T2W, AGH, PGH to PBH lesions, respectively. The peak on the left side of the distribution in NAWM regions becomes smaller with the shift and becomes very small in PBH lesions. Water fraction distributions were essentially the same for all five lesions types, showing narrow distributions of small values.

**Figure 2.**
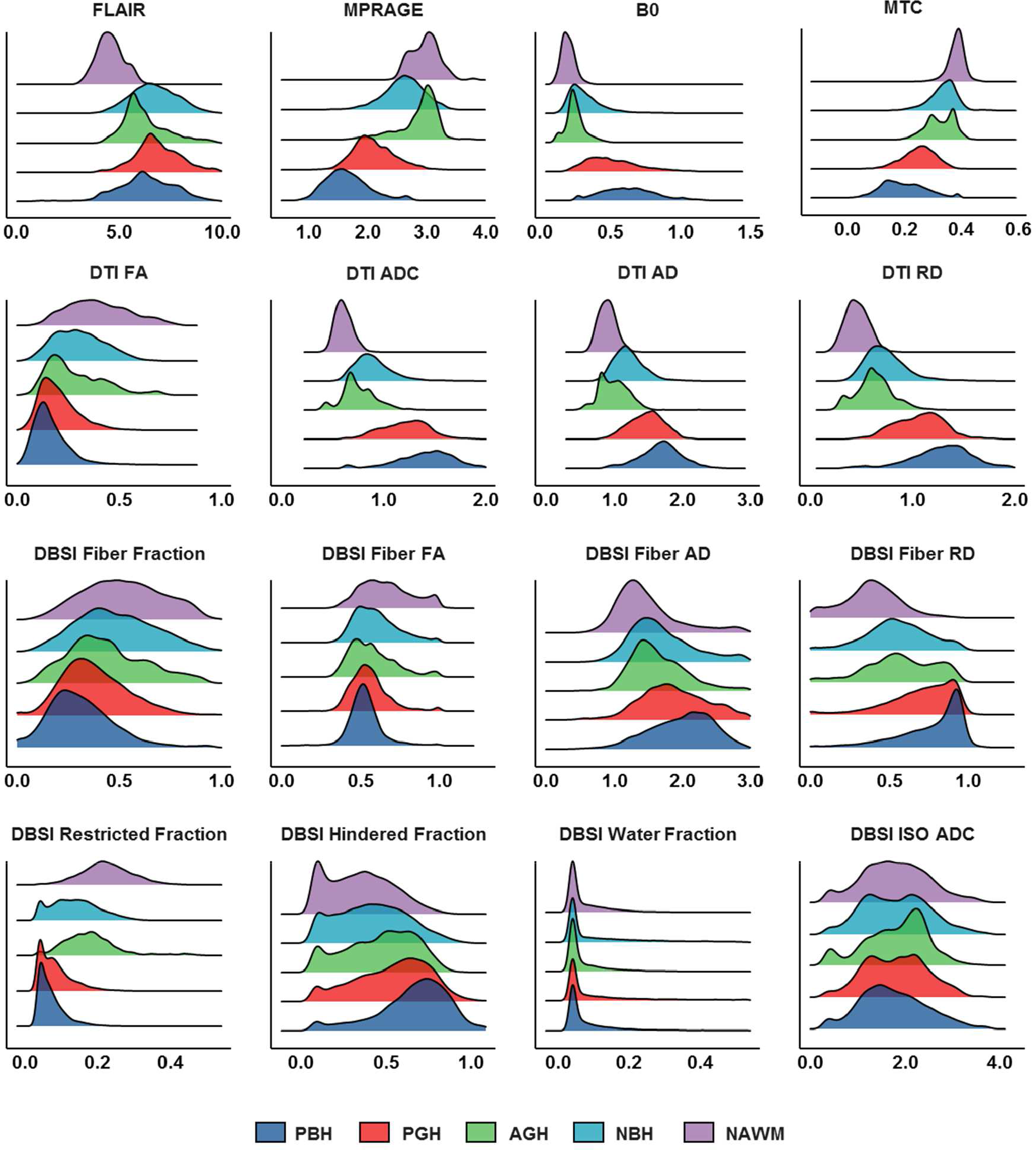
Distribution histograms of different MRI metrics for different MS lesion types. Imaging voxels from persistent black holes (PBH, dark blue), persistent grey holes (PGH, red), acute grey holes (AGH, green), non-black hole lesions (NBH, cyan) and normal appearing white matter (NAWM, purple) were plotted to show the distributions for conventional MRI, MTR, DTI and DBSI metrics.

Overall, different MRI metrics show various distribution patterns among different MS lesion\region types. While some lesion\region types show specific separations, most of the MS lesion\region distributions overlap significantly under conventional MRI. This makes it difficult to accurately distinguish different lesion\region types from each other based solely on MRI features.

### DNN model optimization and validation

DNN model optimization were assessed by comparing overall validation accuracies for DHI model. Neural nets with 9, 10 and 11 hidden layers have smaller standard deviations, while neural nets with much more or much fewer than ten hidden layers have greater standard deviations, indicating better reliability of neural nets with 9, 10, and 11 hidden layers (Fig. 3A). Further, we also assessed models by comparing the number of training epochs and nodes in each hidden layer required for the neural nets to reach 90% validation accuracy. Neural nets with between 100 and 200 nodes per hidden layer produced the best results, which consistently required less than 60 training epochs to achieve a 90% validation accuracy (Fig. 3B). Neural nets with fewer nodes per hidden layer required an increasing number of epochs for training to attain 90% validation accuracy at an exponential rate. In sum, we demonstrated that the neural net with 10 hidden layers and 100 nodes in each layer produces a model with an increasing function and converges to over 90% with minimal standard deviation by 100 training epochs (Fig. 3C).

**Figure 3.**
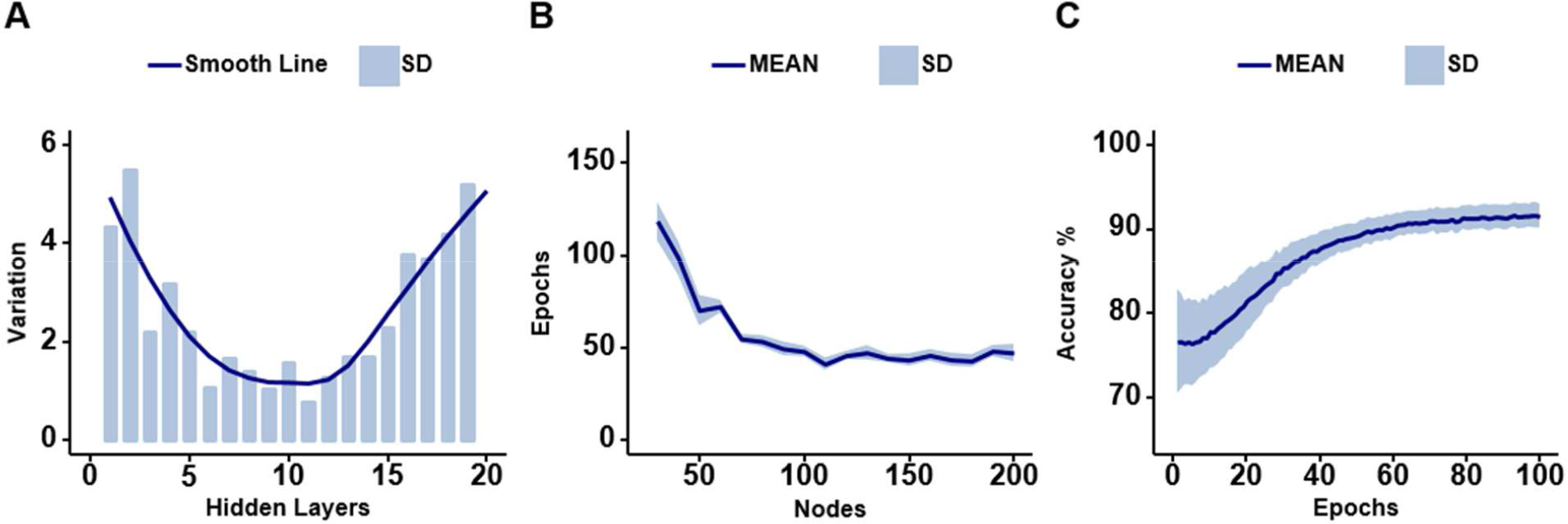
The optimization of DHI model using varying number of hidden layers and nodes. **A**, Neural nets with 1 to 20 hidden layers were tested using 70 repeated random splits of the data for validation accuracy. Total 80% of the data was used to train the DNN, 10% was used for testing and another 10% for validating. Reliability/predictability of neural nets are modelled by standard deviation. Standard deviations of one are shown. All hidden layers tested contain 100 nodes. **B**. The number of epochs required for neural nets to reach 90% validation accuracy are shown. Neural nets with 10 hidden layers and 10 to 200 nodes in each hidden layer were tested. **C**. The optimized neural net of 10 hidden layers each containing 100 nodes was tested on 70 repeated random splits of the data for validation accuracy. The graph shows the validation accuracies over these trials.

### Test Performances of Different DNN Models

DHI model achieved a 93.4% overall concordance with neurologist determinations of all five MS lesions, which outperformed the 78.3% rate from MTR-DNN model, the 80.2% accuracy from DTI-DNN model and the 74.2% accurate prediction from cMRI-DNN model. We used confusion matrices to indicate the discordances between model predictions and neurologist-determined lesion/region types derived from the DHI model (Fig. 4A), the DTI-DNN model (Fig. 4B), the MTR-DNN model (Fig. 4C), and the cMRI-DNN model (Fig. 4D). For one independent test dataset (n=4326), DHI detected PBH, PGH, AGH, T2W, and NAWM with positive prediction rates of 91.3%, 83.4%, 90.1%, 92.3% and 97.9%, respectively, showing very high detection rates.

DTI-DNN performed well in detecting PBH, NBH, and NAWM with true positive rates of 80.1%, 84.8%, and 95.8%, respectively. PGH and AGH were still poorly detected, with a mere 45.7% and 48.1% rates, respectively. DTI-DNN falsely predicted PGH to be PBH (23.9%) or NBH lesion (28.3%). AGH was also mostly false predicted to be T2W lesion (32.2%) and NAWM (16.4%). MTR-DNN model detected NAWM well with a 94% rate. PGH detection was only 63.3% accurate. 17.4% of PGH was incorrectly predicted to be PBH, and another 17.4% of PGH was incorrectly predicted to be NBH lesion. AGH was poorly detected with only a 35% rate. Large portion of AGH was falsely precited to be NBH lesion (38.1%) and NAWM (16.4%). CMRI-DNN model showed good true positive rates on NBH (82.3%) and NAWM (96.1%) lesions. However, this model didn’t perform well on other lesion types. Specifically, PGH were incorrectly predicted to be PBH (24.4%) and NBH (37.8%) lesions. The true prediction rate on PGH was only 35.2%. AGH lesions were also largely incorrectly predicted to be NBH and NAWM with percentages of 55.1% and 38.3%, respectively. The true prediction rate of AGH was only 3.2%.

**Figure 4.**
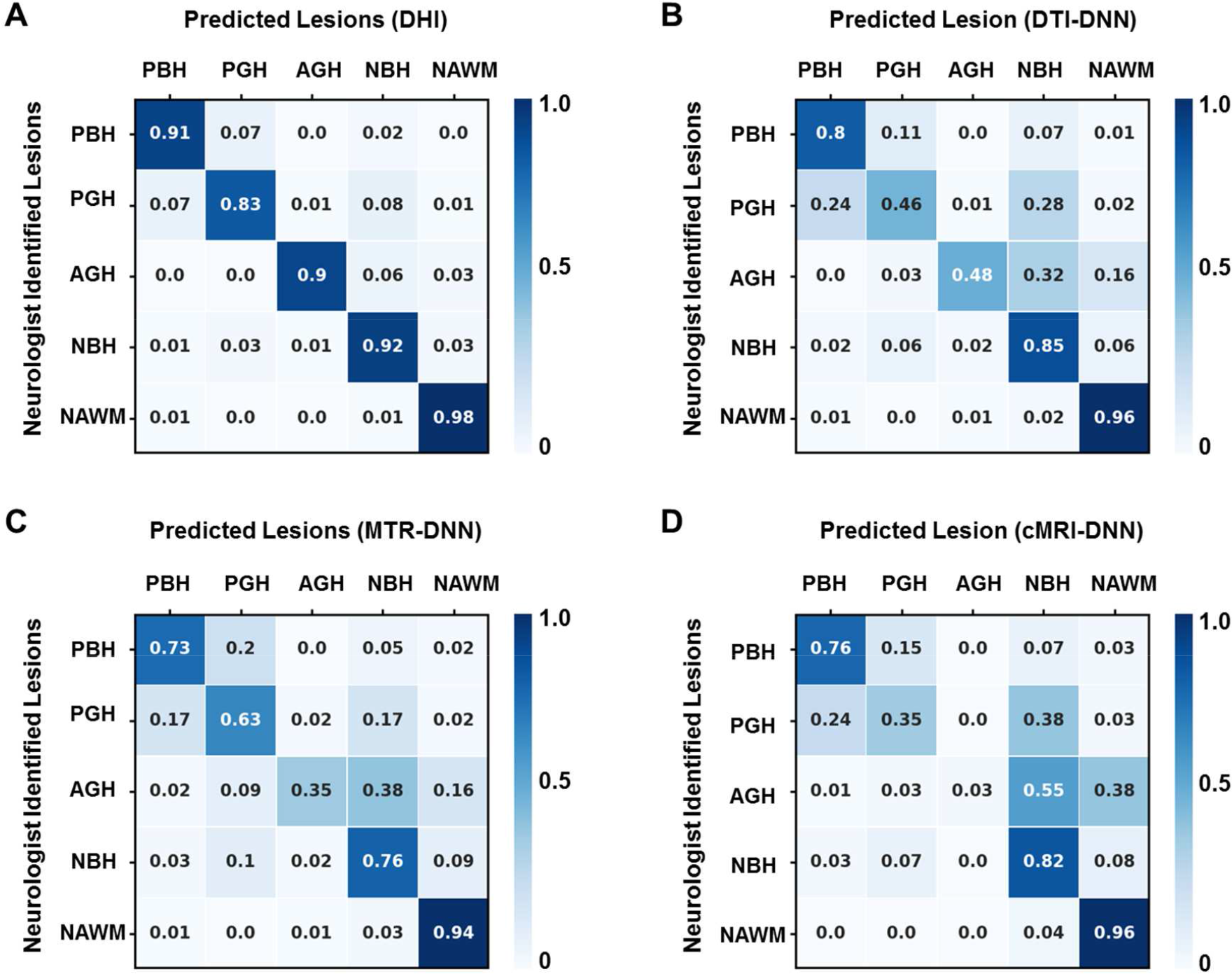
Confusion matrices by DNN trained on various inputs. Rows contain lesion classifications identified by experienced Neurologists (PBH - persistent black hole; PGH - persistent grey hole; AGH - acute grey hole; NBH - non-black hole lesions; NAWM - normal appearing white matter). Columns contain lesion classifications as predicted by different DNN models, including **(A)** DHI (DBSI-DNN model) DHI, **(B)** DTI-DNN model, **(C)** MTR-DNN model and (**D)** cMRI-DNN model.

The one-versus-rest classification strategy was used to calculate ROC and precision-recall curves to test each DNN model’s detection performance on individual lesion/region type. ROC (Fig. 5A) and precision-recall (Fig. 5B) curves from each class of the five lesion/region types were plotted together for comparison. DHI demonstrated the best performances on both ROC and precision-recall analyses, indicating higher ROC AUC and precision-recall AUC values than any other model. DTI-DNN, MTR-DNN and cMRI-DNN showed pretty good performances on ROC analyses, with AUC values for all classes higher than 0.860 (Fig. 5A). However, as ROC analysis is insensitive to class imbalance, the ROC AUC values could overestimate the model performances. Precision-recall curve could effectively address this issue and provide complimentary informaton to ROC. These three models didn’t perform as well on precision-recall analyses. For example, the precision-recall AUC values for PGH and AGH in these three models were all lower than 0.650 (Fig. 5B). We used bootstrap method with 1000 itertations to calculate ROC AUC, sensitivity and specificity values for DHI model and summarize the results in Table 2. We also calculated F_1_-score for all the five classes. In particular, we can see the great performance of DHI in detecting PBH (AUC: 0.991 (95% CI: 0.989-0.994); F_1_-score: 0.923), PGH (AUC: 0.977 (95% CI: 0.971-0.982); F_1_-score: 0.823) and AGH (AUC: 0.987 (95% CI: 0.980-0.992); F_1_-score: 0.887).

**Table 2.**
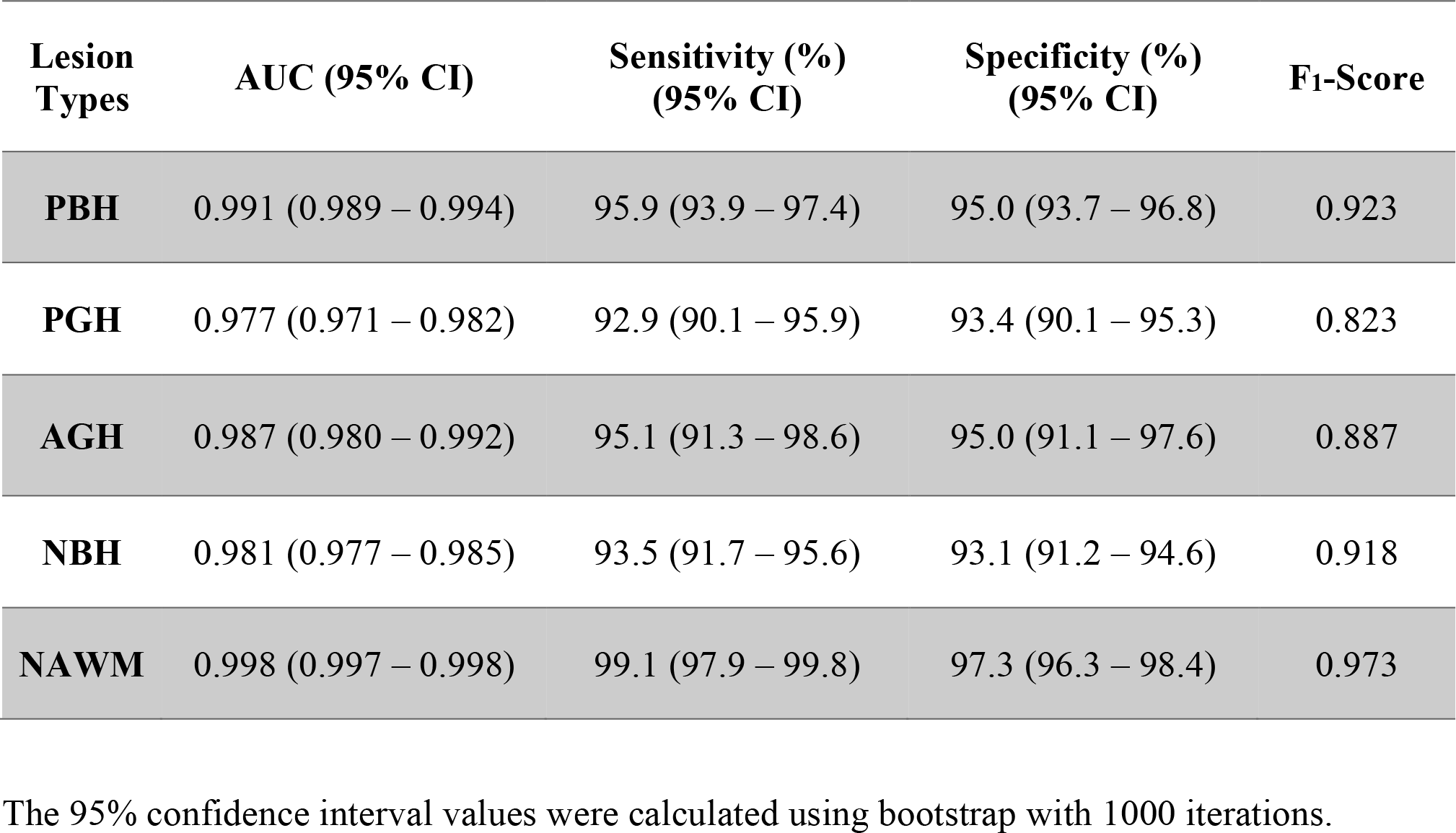
Diagnostic performances of DHI models.

**Figure 5.**
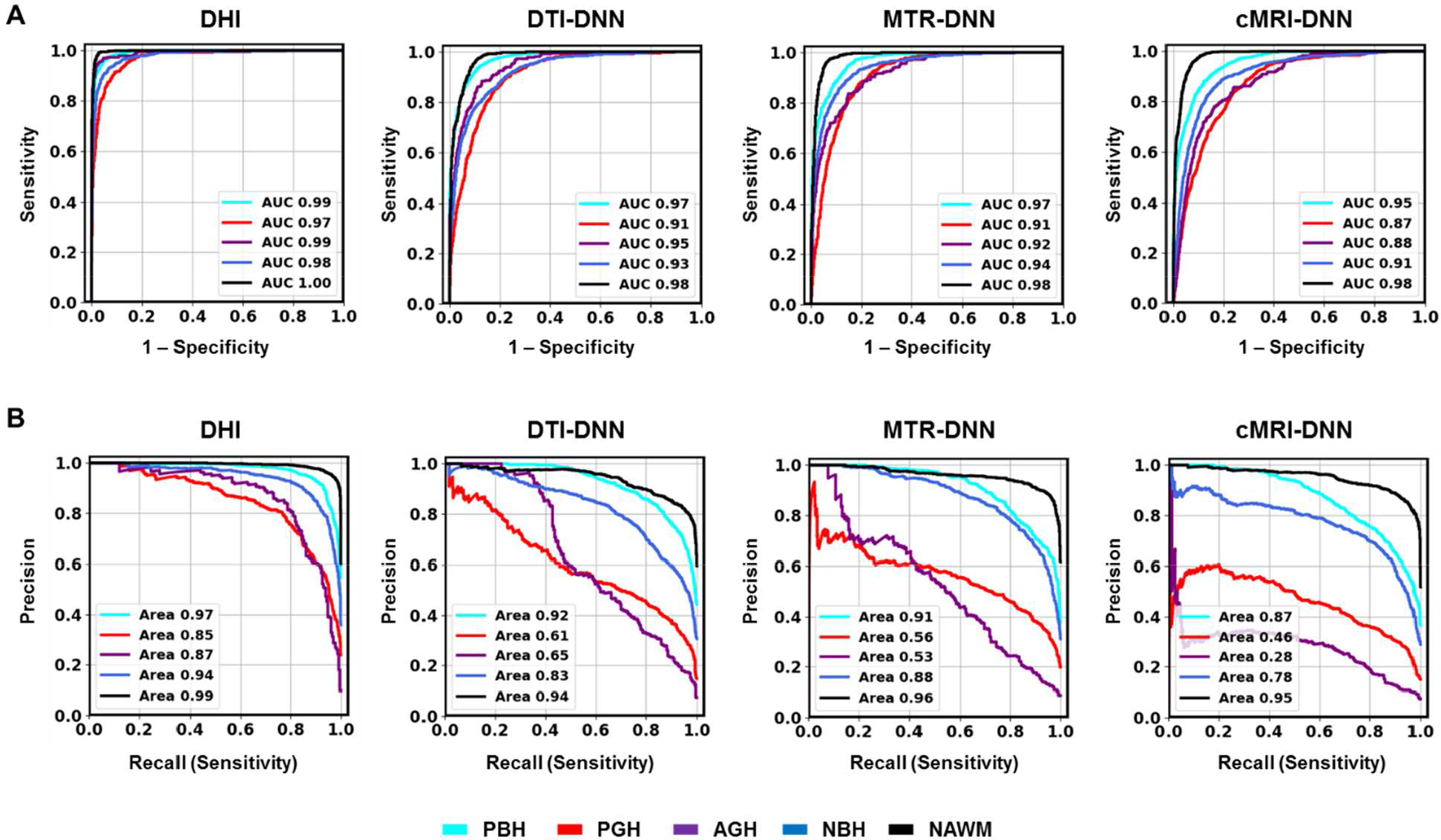
ROC and precision-recall curves. **A.** Examples of ROC curves calculated on an independent test set (n=4,327) by four different models of DHI, DTI, MTR and cMRI. **B**. Examples of precision-recall curves calculated by DHI, DTI, MTR and cMRI models. DBSI showed the greatest performances on both ROC and precision-recall curves for all the five different lesions compared with other models. Class labels are as follows: Cyan, PBH. Red, PGH. Purple, AGH. Blue, T2W. Black, NAWM.

## Discussion

We have developed a novel diffusion basis spectrum imaging (DBSI) and demonstrated its capability to quantitatively characterize the pathologies underlying MRI lesions in MS patients and post-mortem specimens. Our main motivation was to examine whether the inclusion of DBSI metrics in our optimized deep neural network model will increase classification accuracies for identifying MRI lesions when compared to other established methods. Our results have clearly demonstrated that the use of DBSI, T1WI and T2WI as neural network inputs produces more accurate classification results. The confusion matrices indicated that models trained on DBSI metrics have higher positive prediction rates for each of the five lesion types than the other three commonly used metric sets for training. The ROC and precision-recall curves clearly show that the DHI model has greater overall classfication performances for all 5 lesion and tissue types than the other 3 commonly used metric sets. Furthermore, a wide range of diagnostic performance metrics show that predictions from the DHI model outshine the other three models trained on commonly used metric sets.

MRI has played a vital role in the diagnosis and management of MS over the past decades^19^. However, conventional T1W and T2W brain imaging techniques do not correlate well with MS pathologies. One of the main reasons is the complex pathologic heterogeneity of MS lesions^20,21^. Furthermore, conventional T1W and T2W imaging contrasts vary from scan to scan and are not quantitative, as they depend not only on the MR characteristics of brain tissue but also the scanner vendors, magnet strength, and pulse sequences. Recently, DTI has been widely applied in brain imaging for multiple central nervous system (CNS) disorder^22-24^, as it has been shown to correlate with axonal injury and demyelination^25-27^. However, DTI is based on a single tensor Gaussian diffusion model, which makes it inadequate in resolving co-existing complicated pathologies such as neuroinflammation^28^. To address this issue, DBSI adopts a computationally novel model that separates isotropic and anisotropic components from each other in imaging voxels ^9,29,30^. Isotropic diffusion is believed to reflect inflammatory components (cells, edema), as well as intrinsic cells and extra-cellular space, while anisotropic components could reflect the integrity of axon fibers and myelin. DBSI enables the detections and quantifications of these histological components and associated pathological changes. This has been shown in the EAE mice model and human specimen, where we demonstrated that the DBSI-derived metrics of axial diffusivity, radial diffusivity, and restricted isotropic diffusion fraction had significant correlations with corresponding immunohistochemical stains^11,31^. DBSI-derived metrics have also been shown to reflect various specific components of the CNS pathology in patients with MS^10,11^. These correlations strongly support the utility of DBSI to accurately measure axonal injury, demyelination as well as increased inflammatory cellularity. Those pathology-specific metrics from DBSI enable more accurate classifications of various types of MS lesions.

Different MS lesion types are associated with different CNS pathology and patient outcomes. The pathologic correlation of PBH lesions is the severe axon loss and matrix destruction^32-34^. PBHs are believed to exhibit more axon loss than other types of MS lesions neuropathologically^33^. The amount of PBHs present and the volumes of those PBHs showed positive correlations with increased MS-induced disabilities^35,36^. Additionally, several other studies have shown that PBH numbers or volumes correlate with worse clinical test scores^35^. This shows that PBHs have significant relevance to clinical outcomes and disease progression. Compared to PBHs, PGHs reflect a lower degree of axonal loss. Comparisons of imaging and neuropathology in over 100 MS lesions found with a certain degree of hypointensity was strongly associated with axonal density^37^. MS lesion burden has often been reported as the sum of lesion volumes, but the degree of tissue destruction may vary between lesions^38^. A quantitative method to distinguish the degree of hypointensity in individual MS lesions could improve patient monitoring, allow for better correlations to clinical measures, and be useful as an outcome measure in clinical trials of potential reparative therapies^37^. In contrast with PBH and PGH lesions (which are associated with axonal loss), ABH and AGH lesions are likely caused by inflammation and edema, since most ABH/AGH lesions resolve within months of contrast resolution^22^. Although these lesion types reveal different pathological characteristics, they all show hypointensity in pre-contrast T1W images. Clinically it is challenging to distinguish these lesions solely by expert raters consistently. In contrast, our DNN based tool could automatically and accurately predict different lesion types, showing great promise to effectively and efficiently assist clinicians.

MS lesion identification and segmentation based on manual tracings of structural boundaries and signal intensity remained the most commonly used MS diagnostic method. This method not only requires experienced radiologists or neurologists, but is also time-consuming, expensive, and subject to operator bias, which often causes considerable inter- and intra-rater variability^39^. In addition, it is difficult for a human expert to combine information from various slices and channels when multispectral MRI data are examined^40^. Computer-aided analysis can assist in identifying brain structures and lesions, extract quantitative and qualitative properties and evaluate their progress over time. Automatic methods of image segmentation and classification have been investigated extensively. Recently, supervised machine learning methods such as logistic regression^41^, k-nearest neighbors^42^, support vector machine^43^ as well as deep convolutional neural network (CNN)^44^ have been explored to segment MS lesions from normal brain regions. However, classifications for various types of MS lesions using deep neural networks have not been reported. Therefore, we constructed a DNN model employing DBSI metrics that showed high capability in distinguishing different MS lesion types.

There are some limitations of this study. First, the subject number is relatively small with only 38 MS patients included in our study cohort. However, a total of 43,261 imaging voxels from 499 MS lesions were identified by expert neurologist from three subtypes of MS patients, which could effectively remedy the small cohort size of the study. Additionally, DBSI derived metrics were modeled and calculated on a single voxel basis, which would address the heterogeneity of MS lesions. Second, the data distribution was imbalanced among different lesion\region types, which could compromise the performance of a DNN model. While the class imbalance is a common issue in most clinical studies, DBSI provided pathology-specific features that significantly improve the overall performance for the DNN model.

## Conclusions

DHI significantly improves the accuracy of MS detection and classification for five representative MS lesion types, especially when the overall prediction accuracy is 91.2%, which significantly outperformed other models. The improved DHI framework could greatly assist neurologists and neuroradiologists in making clinical decisions. The efficiency and effectiveness of this DNN model demonstrates great promise for clinical application and the potential to become a marker of lesion severity and possibly lesion recovery for clinical trials and in practice. Additional longitudinal studies with larger cohorts, different scanners, and multiple centers are imperative for further explore the possibilities of applying DHI on a broader scope; this DHI may aid in assesing therapeutic efficacy with regards to axonal degeneration in trials.

## Data Availability

The datasets and codes from this study are available from the corresponding author on reasonable request.

## Author Contributions

Z.Y., P.S., A.H.C. and S.S.-K. designed the study, supervised the experiments and wrote the manuscript. Z.Y., A.W., A.G., X.N., J.L. and G.A. performed the experimental work and analysis and refned the manuscript.

